# Social Distancing and Personal Protective Measures Decrease Influenza Morbidity and Mortality

**DOI:** 10.1101/2020.04.17.20070102

**Authors:** Grant Young, Xiaohua Peng, Andre Rebeza, Santos Bermejo, Chang De, Lokesh Sharma, Charles S. Dela Cruz

**Author notes:** equal contribution as first authors (Grant Young, Xiaohua Peng). equal contribution as senior authors (Lokesh Sharma, Charles S. Dela Cruz). Corresponding authors: Lokesh Sharma and Charles S. Dela Cruz.

## Abstract

Seasonal influenza (flu) is an underappreciated source of disease morbidity and mortality worldwide. While vaccination remains the cornerstone of influenza prevention, common measures practiced during the COVID-19 pandemic such as social distancing, the use of protective face masks, and frequent hand washing are rarely utilized during flu season. In this investigation, we examined the effect of these preventative measures in decreasing influenza burden this year. We examined three countries with major COVID-19 outbreaks i.e. China, Italy and the United States, and compared the flu activity this year to the average of the last 4 years (2015-2019). We found that this year in China and Italy, there was a significantly steeper decline of flu cases than average, which correlated with an increase in positive COVID-19 case reports in those countries. These "averted" cases can be translated into a substantial decrease in morbidity and mortality. As such, we conclude that the current COVID-19 pandemic is a reminder that behavioral measures can decrease the burden of communicable respiratory infections, and these measures should be adopted to an extent during normal influenza season.

## Introduction

Seasonal influenza (flu) is a leading cause of disease burden worldwide. Each year, the flu epidemic results in up to 5 million severe cases and 650,000 deaths, which are staggering numbers that exceed those of any other infectious disease in high income countries.^1^ Influenza season varies depending on the geographical season. For example, peak influenza activity is observed between October and March in China while in the United States, influenza season starts in October, peaks in February, and ends in May.

Vaccination is the foremost current approach in influenza prevention. However, seasonal vaccination is only partially effective in disease prevention and efficacy depends on the accurate prediction of the circulating strains. Other measures such as social distancing, the use of protective face masks, and frequent hand washing are rarely practiced during influenza season. However, during the current outbreak of COVID-19, these protective steps were applied to reduce person to person transmission of virus. It is clear that these additional initiatives have helped to curb the number of cases, especially in countries such as China and South Korea.

Therefore, it is reasonable to suspect that implementing these measures to a less extreme extent during the normal influenza season may also help to diminish influenza spread, subsequent hospitalizations, and mortalities. This strategy may be particularly relevant in protecting populations who are at higher risk of contracting severe influenza disease. In this study, we investigated the role of current preventive measures taken in response to COVID-19 outbreak in decreasing the cases of influenza virus and associated mortality.

## Methodology

Influenza activity and the numbers were obtained using data from the World Health Organization (WHO) website https://www.who.int/influenza/gisrs_laboratory/flunet/en/. The new cases of COVID-19 were also sourced from the WHO https://experience.arcgis.com/experience/685d0ace521648f8a5beeeee1b9125cd. Data were obtained from three different countries which had major COVID-19 outbreaks i.e. China, Italy and United States of America (USA). We first compared the activity of influenza this year to the average of the last four years (2015-2020), using China as an example (Fig 1A). Influenza activity was defined each week as local outbreak (1), regional outbreak (2) or widespread outbreak (3). Subsequently, we devised a method to account for two considerations: 1) the timing of flu season varies each year and also among the countries, and 2) the number of positive flu cases is different each year. To address the first concern, for each flu season we arbitrarily assigned the peak (the week in which the most cases of flu were diagnosed) as week 0. Weeks before the peak of the flu season were assigned (-) values and weeks after the peak were assigned (+) values. Second, we standardized the variations in positive cases for each flu season by dividing the number of positive flu cases in a given week by the number of positive cases in week 0. The resulting “standardized fraction” is plotted on the left Y-axis of Figures 1B-D. The ratio of the standardized fractions in 2019/2020 compared to the average for 2015-2019 was plotted on the left Y-axis (grey) of Figures 2A-B. The right y-axis (blue) shows the number of flu cases averted by COVID-19 policies, calculated by multiplying (1-ratio) with the average numbers of (2015-2019) number of cases for that week, multiplied by 140. The 140 comes from https://wwwnc.cdc.gov/eid/article/15/12/09-1413_article, an article cited by the CDC to approximate actual flu cases under the assumption that positive tested cases are a gross underestimate.^2^

**Figure 1:**
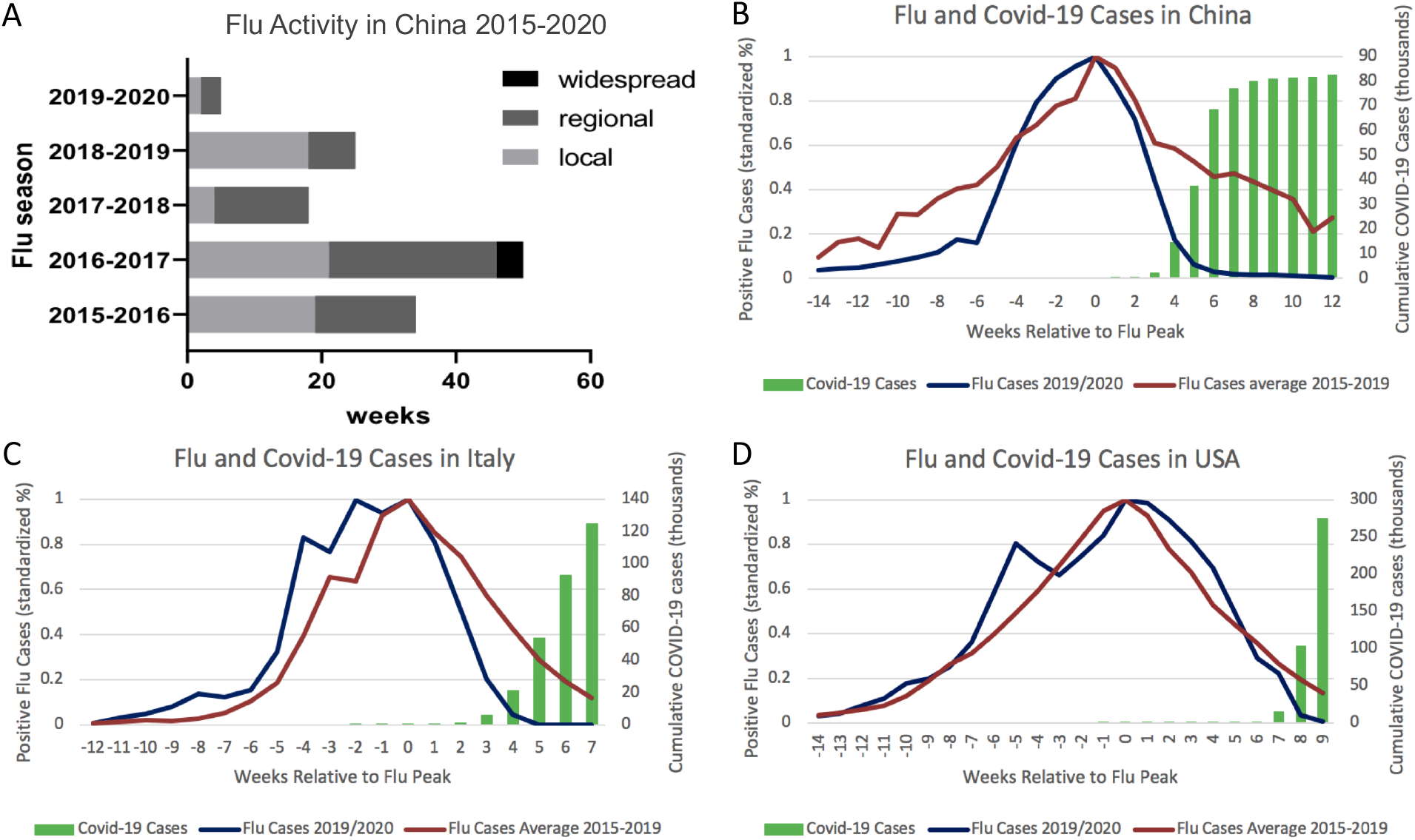
A, Flu activity in China from 2015 to 2020. Local, regional and widespread outbreaks are defined by FluID_InfluenzaEPIform. B-D, Flu cases in China, Italy, and the United States in 2019/2020 (blue) compared to the average for 2015-2019 flu seasons (red). The x-axis shows weeks leading up to and following the peak at week 0. Flu cases are shown on the left y-axis as a standardized fraction. Covid-19 cases (green) are shown on the right y-axis.

**Figure 2:**
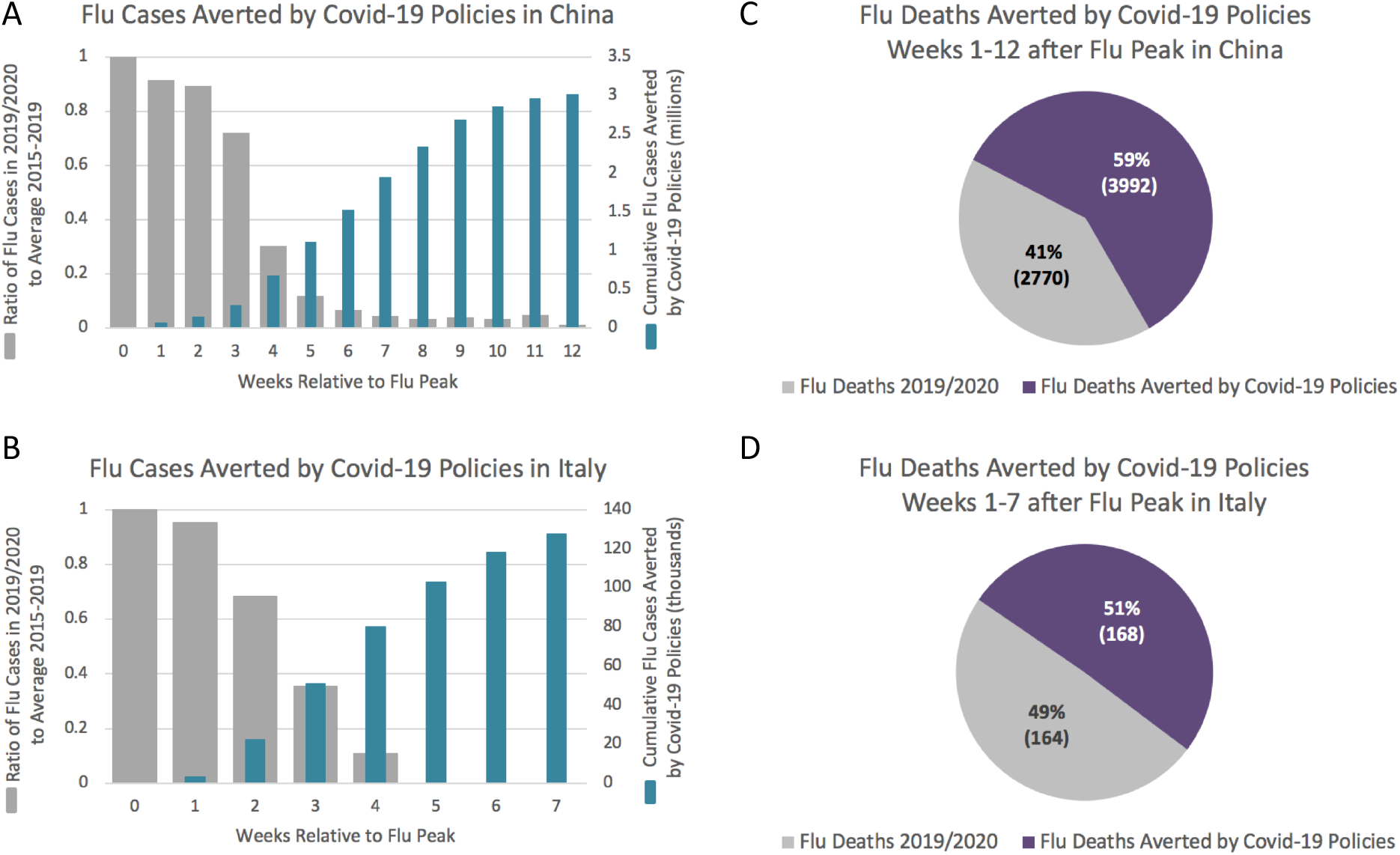
A-B, Flu cases averted by Covid-19 policies in China and Italy. The left y-axis (grey) is the ratio of standardized flu cases in 2019/2020 compared to average. The right y-axis (blue) shows cumulative flu cases “averted.” C-D, Flu deaths averted by Covid-19 policies in China and Italy. Purple represents flu deaths averted in 2019/2020 compared to average. Grey represents flu deaths that still occurred despite social distancing.

In Figures 2C-D, the number of flu attributed deaths averted by COVID-19 policies was calculated using the percentage of cases averted (1-ratio of grey bar in figures 2A-B), multiplied by the average number of cases of flu diagnosed that week (2015-2019 average), multiplied by 140, multiplied by the mortality (.132%).^3^ This was done for each week in the given period of time, then the weeks were summed to give the number of total deaths averted for that timeframe.

## Results

### Influenza cases decreased significantly due to COVID-19 outbreaks in China and Italy

First, we studied how the presence of COVID-19 affected influenza activity. In China, it is evident that flu activity in prior years (2015-2019) had a significantly longer duration than the current season (Fig. 1A). In flu seasons from 2015-2019, the influenza activity remained at level 2 for 15.33±9.07 weeks and at level 1 for approximately 14.33±9.07 weeks (means + SD). The current season only had brief level 1 activity for 2 weeks and level 2 activity for 3 weeks. We further explored this phenomenon using actual records of positive flu cases in China, Italy and USA (Fig. 1B-D). This year, in China and Italy, we observed a steep decline in influenza cases following the peak of activity at week 0 compared to the average year (2015-2019). Strikingly, this decline in influenza inversely correlated with an increase in positive COVID-19 cases. This trend is most obvious in China, followed by Italy, while there was a slight decrease in flu cases this year compared to average in the United States.

### Estimation of flu cases and mortality averted by COVID-19 outbreak

Next, we sought to estimate the number of influenza-attributed cases and deaths that were averted by protective measures aimed to stem the COVID-19 outbreak in China and Italy. We first used a ratio to more clearly visualize that compared to the average (2015-2019), the number of flu cases this year declined steeply after the peak of the season both in China and Italy (grey bars, Fig. 2A-B). The blue bars show that a significant number of influenza cases in China (n = 3.02 million, 12 weeks after peak) and in Italy (n = 127 thousand, 7 weeks after peak) were averted this year. Translating these cases to fatalities averted, our data showed that 3992 (59%) of estimated deaths in China and 168 (51%) of estimated deaths in Italy were avoided this flu season (Fig. 2C-D). Taken together, these data show that behavioral changes can prevent a significant number of flu cases.

## Discussion

Seasonal influenza has been an underappreciated threat in high-income countries for many decades. In fact, influenza frequently outpaces all other infectious diseases in terms of cases and deaths on a yearly basis. Despite its immense disease burden, the effort to contain the influenza virus by limiting human to human transmission remains minimal. The current outbreak of COVID-19 and policy measures taken to prevent the spread of the SARS-CoV-2 virus have provided us with an important opportunity to investigate how restricting human to human transmission can affect influenza associated morbidity and mortality.

First, we show that the duration of influenza activity this year was significantly shortened by the current COVID-19 outbreak in China. The sharp decline in flu activity was correlated with rising cases of COVID-19 and subsequent social distancing measures. It is plausible that fewer influenza tests were performed this year due to COVID-19, but this seems unlikely. Studies have reported testing for influenza A and B in most of the suspected COVID-19 cases that presented with flu like symptoms.^4^ It is also unlikely that viral interference, a known mechanism in which an immune response to one viral infection prevents a second viral response, contributed to limited influenza spread, as co-infection with influenza and COVID-19 has been reported.^5^ The shortened activity of influenza was associated with a decrease in the number of cases, a phenomenon evident in China and Italy but less so in USA. The lack of decline in US cases may be a function of late implementation of social distancing measures, or the fact that the flu season in the US was already nearing its end when these measures were adopted.

The estimated number of “lives saved” (59%, 3992) by social distancing policies in China seems to be comparable to the reported number of COVID-19 attributed mortalities (3338), as of April 5^th^.^6^ In Italy, a similar percentage (51%) of lives were “saved,” but the actual number (168) was much lower (Fig. 2C-D). We did not calculate this number for the US as we saw minimal effect on the influenza disease burden this year. These data show that quarantine and other social distancing measures are only effective when implemented in a timely manner.

In conclusion, we recommend that some social distancing should be adopted normally during influenza season, especially for those in elderly and other high-risk population. The current COVID-19 pandemic comes as a great reminder that simple behavioral measures such as social distancing and effective hand washing can greatly decrease the burden of communicable respiratory infections.

## Data Availability

Publicly available data
https://www.who.int/influenza/gisrs_laboratory/flunet/en/, https://experience.arcgis.com/experience/685d0ace521648f8a5beeeee1b9125cd

https://experience.arcgis.com/experience/685d0ace521648f8a5beeeee1b9125cd

